# Study protocol Improving inpatient GHB detoxification with baclofen: a proof of concept and dose-finding study

**DOI:** 10.1101/2025.08.05.25333021

**Authors:** A.M.L. Wood, H. Beurmanjer, B.A.G. Dijkstra, A.F.A. Schellekens

## Abstract

**Background:** Gamma-hydroxybutyric acid (GHB) use disorder (GUD) is well known for its severe withdrawal syndrome. Currently, tapering with pharmaceutical GHB is the preferred option to mitigate withdrawal. The gamma-aminobutyric acid (GABA)-B receptor agonist baclofen could improve GHB detoxification, due to its longer half-life than pharmaceutical GHB and specific GABA-B receptor activation. However, clinical data on baclofen’s effectiveness to manage GHB withdrawal is limited. We aim to explore potential benefits of baclofen as an add-on to pharmaceutical GHB tapering in patients with GUD.

**Methods:** This prospective, non-randomized, clinical trial consists of two phases: 1) a proof-of-concept phase (n=10) and 2) a dose-finding phase (n= 18). Primary objective of the proof-of-concept phase is to assess whether baclofen add-on therapy reduces the need for pharmaceutical GHB during inpatient detoxification. Safety and feasibility will also be assessed. Aim of the dose-finding phase is to explore the optimal baclofen dosage when used as add-on therapy during inpatient GHB detoxification. Criteria to start the second phase are that baclofen add-on therapy leads to a dose reduction of pharmaceutical GHB without severe side effects. GHB withdrawal symptoms and baclofen side effects will be monitored during GHB detoxification, using questionnaires.

**Discussion:** We expect that baclofen add-on therapy will reduce the need for pharmaceutical GHB during inpatient GHB detoxification. This will likely result in a less intensive detoxification process for patients and will improve the overall feasibility of GHB detoxification. If the outcome of the current phase II trial is positive, either a replication phase II trial with a randomized blinded design or a lager phase III trial, would be a logical follow-up study to confirm the safety, feasibility and efficacy of baclofen add-on therapy during inpatient GHB detoxification.

**Trial registration:** This study protocol is approved by the Medical Ethical Research Committee Oost-Nederland and Central Committee on Research Involving Human Subjects. The study is registered in the Clinical Trial Information System under EU CT-number: 2023-506167-34-02.

## Background

Gamma-hydroxybutyric acid (GHB) is an addictive substance, used recreationally for its euphoric, relaxing and sexually stimulating effects in combination with a boosted self-confidence [1–4]. However, repeated recreational use of GHB can escalate quickly and lead to tolerance and dependence within weeks [1,5,6]. Like other substance use disorders (SUD), GHB use disorder (GUD) is characterized by craving, lack of control over substance intake, and physical dependence [7]. GHB use disorder (GUD) is associated with high relapse rates of 50% within three months after detoxification and high re-admission rates [8–10]. When chronic GHB use is suddenly stopped or tapered off too quickly, a severe withdrawal syndrome can occur, characterized by tremors, agitation, anxiety, hallucinations and hemodynamic instability [2,11,12]. When withdrawal symptoms are not managed properly there is the risk of developing a delirium or psychosis [13].

The occurrence of withdrawal symptoms after abrupt cessation of GHB use is based on a complex interaction of several neurobiological mechanisms in which glutamatergic overactivation and gamma-aminobutyric acid (GABA)-ergic under activation are thought to play key roles [5]. GHB mainly acts via a bidirectional effect on GABA receptors and via activation of the GHB-receptor. At low doses GHB primarily affects the high-affinity metabotropic GHB receptor, leading to an increase in glutamatergic signaling and a decrease in GABA-ergic signaling [5,14,15]. At high doses GHB activates the low-affinity GABA-B receptor, decreasing glutamatergic signaling and increasing GABA-ergic signaling [5,16].

Currently, there are two commonly used methods for detoxification of patients with GUD: benzodiazepine (BZD) tapering and pharmaceutical GHB tapering [13,17,18]. BZDs increase the sensitivity for GABA, by acting on the GABA-A receptors. BZDs are widely available in medical settings and low in costs. However, several studies report that BZDs as stand-alone therapy are not sufficiently effective to prevent and treat GHB withdrawal [5,19]. Despite high doses of BZD, severe withdrawal symptoms, such as delirium and psychosis, are often described [5,20]. Pharmaceutical GHB acts at the same receptors as illicit GHB [5,15]. Pharmaceutical GHB-assisted detoxification has been shown to be safe, with less severe withdrawal and less complications than BZD tapering [13,17,18].

One of the main disadvantages of pharmaceutical GHB is the short half-life (20-50 minutes), requiring frequent dosing up to 12 times a day, every 2-3 hour, even at night. Furthermore, patients have to continue using their substance (GHB) during detoxification, which has been suggested to reinforce GHB use, maintaining the addiction cycle and potentially increasing the risk of relapse [13]. This highlights the need for safe alternatives for tapering with BZDs or pharma GHB with a long-acting compound.

Given the partially overlapping profile with GHB, the GABA-B receptor agonist baclofen might be of interest to improve the GHB detoxification process. After administration, baclofen reaches peak plasma concentrations within 2 hours, with a half-time ranging from 2-6 hours [21]. This provides more stable plasma levels and subsequent GABA-B receptor activation with less frequent dosing compared to GHB [20,22–25]. Preclinical studies have shown that baclofen reduced GHB self-administration and GHB-seeking behaviour in mice [26].

Several case reports suggest baclofen might be an effective add-on treatment to BZD’s [22,25,27–29] or stand-alone treatment [30] for GHB withdrawal symptoms. Most studies report low doses of baclofen ranging between 30-80mg/day. However, the use of baclofen in higher doses has also been studied (120-270mg/day) and appears to be safe, with only mild adverse events such as fatigue [30– 32]. Similarly, in the review of Leung et al. (2006) describing the spectrum of toxicity of baclofen in overdose and the dose-related clinical effects, baclofen doses up to 200mg are not associated with adverse effects, such as delirium or coma. Furthermore, baclofen appears popular within the online self-medication community to manage GHB withdrawal or to help increasing time between GHB doses overnight, using a wide variation in dosages [22].

Although the current literature on baclofen for detoxification of GUD seems promising, no clinical trials have yet been conducted on the effectiveness of baclofen as add-on or stand-alone therapy to prevent and treat GHB withdrawal symptoms during the detoxification phase. Data on the optimal dosage for GHB detoxification are lacking. Overall aim of this study is to explore the value of baclofen as add-on therapy during inpatient GHB detoxification with pharmaceutical GHB.

## Objectives and research questions

This clinical trial consists of two phases, both with specific objectives.

### Phase I: proof-of-concept

Primary objective is to assess whether the baclofen add-on therapy reduces the need for pharmaceutical GHB during inpatient detoxification in patients with GUD. Secondary objective is to assess the safety of baclofen add-on therapy during inpatient detoxification in patients with GUD.

#### Primary research question

1.1 Does addition of baclofen to the regular treatment procedure result in less pharmaceutical GHB needed to counteract withdrawal during inpatient detoxification in patients with GUD?

#### Secondary research question

1.2 Which side effects and/or adverse events are reported at the prescribed baclofen dosages during inpatient detoxification in patients with GUD?

### Phase II: Dose-finding phase

Primary objective of the dose-finding phase is to explore the optimal baclofen dosage when used as add-on therapy during inpatient GHB detoxification. Secondary objective is to assess the safety of baclofen add-on therapy during inpatient detoxification in patients with GUD.

#### Primary research questions

2.1 What is the overall impact of baclofen add-on therapy on subjective and objective withdrawal symptoms and craving during inpatient detoxification in patients with GUD?

#### Secondary research question

2.2 What is the impact of different doses of baclofen on the need for pharmaceutical GHB, subjective and objective withdrawal and craving during inpatient detoxification in patients with GUD?
2.3 Which side effects and/or adverse events are reported at the prescribed baclofen dosages during inpatient detoxification in patients with GUD?

## Methods

### Study design

This prospective, non-randomized, phase II clinical trial consists of two phases: 1) a proof-of-concept phase (n=10) and 2) a dose-finding phase (n= 18). Criteria for the transition to the second phase (dose-finding) are that baclofen add-on therapy to treatment as usual (TAU) leads to a reduction in the need for pharmaceutical GHB during inpatient detoxification without severe side effects. For an overview of the study design, see Figure 1.

**Figure 1.**
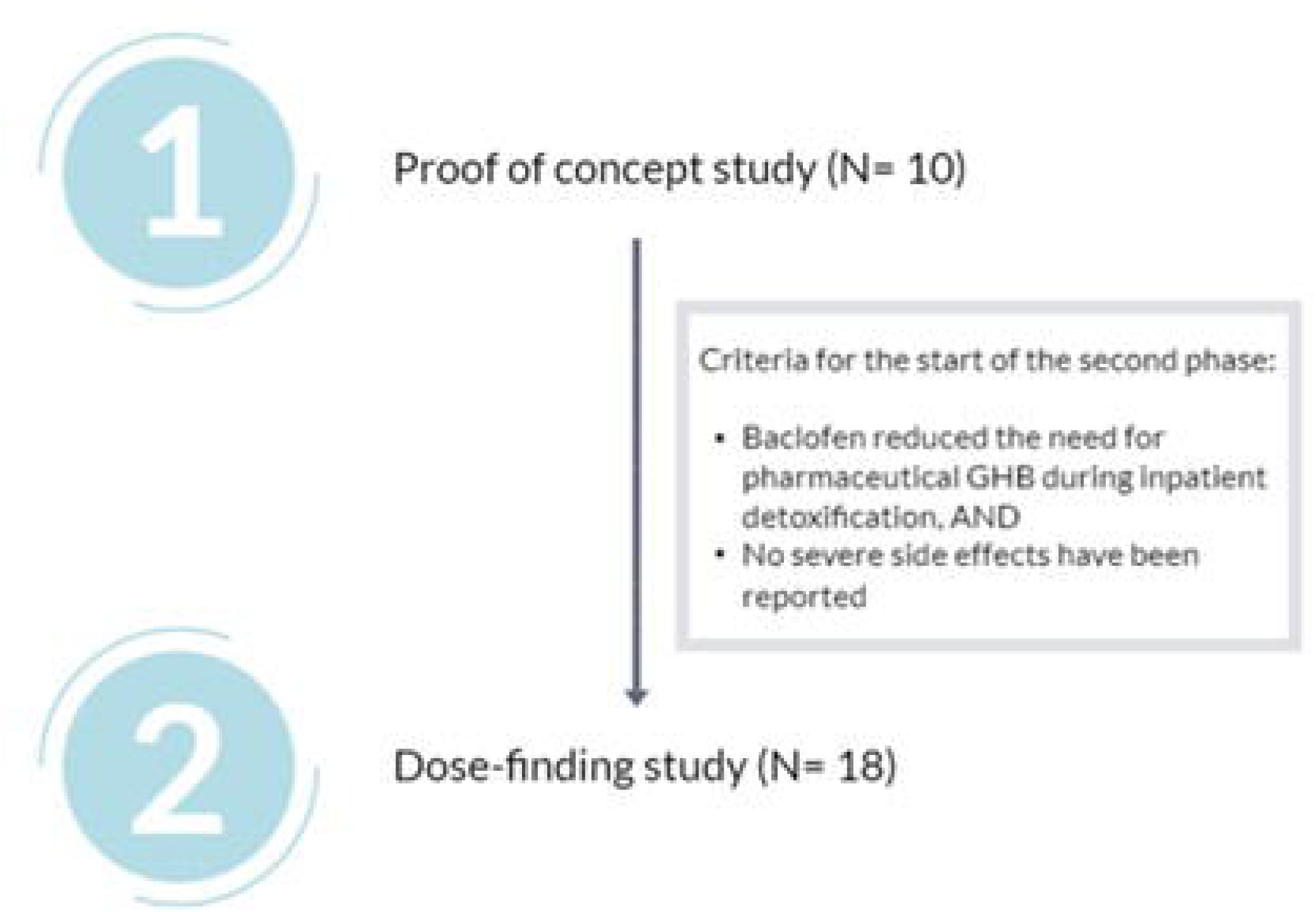
overview of the study design

To assess whether the baclofen add-on therapy reduces the need for pharmaceutical GHB during inpatient detoxification in patients with GUD (objective 1, proof-of-concept phase), a reference group that received pharmaceutical GHB only (TAU) will be used. For the reference group, we will use existing data from two multicenter observational cohort studies of patients with GUD [12,13]. The use of this existing database will lead to a reduction in the number of participants required for the study and will increase the feasibility of the study. The dose-finding phase is an observational study, for which no reference group is required.

### Participants

#### Inclusion criteria

In order to be eligible to participate in this study, a participant must meet all of the following criteria: diagnosed with GUD according to the DSM-5-TR criteria of substance use disorder (1), indication for intramural GHB detoxification (2), GHB use > 50ml/day (3), and > 18 years old (4).

#### Exclusion criteria

A potential participant who meets any of the following criteria will be excluded from participation in this study: 1) having acute psychiatric co-morbidity that requires immediate medical attention and interferes with the execution of the study, such as mania, delirium and psychosis; 2) having physical contra-indications for baclofen: liver problems, renal impairment, heart failure, urine retention, seizure disorder, Parkinson’s disease, and pregnancy; 3) a body weight <55kg; 4) insufficient understanding of the Dutch language; 5) GHB use > 120ml/day, and 6) comorbid benzodiazepine use > 30 mg diazepam equivalent per day.

### Treatment methods

#### Treatment as usual (TAU)

Patients will be treated with pharmaceutical GHB using a titration and tapering method. Patients will start with 70% of their daily street GHB dose, divided by eight to match the dosing interval of 3 hours. Next, the GHB dose is titrated up in case of withdrawal and down in case of sedation. When the patient is stabilized, the tapering phase starts in which the pharmaceutical GHB (500mg/ml) is lowered with 0,6ml/gift/day. For a more detailed description of the titration and tapering (DeTiTap) method, see Dijkstra et al., 2017.

#### Baclofen proof-of-concept study

The first dose of baclofen is given when the patient experiences the first symptoms of withdrawal (see table 1) or when the time for the patient’s usual GHB dose interval has elapsed. As with TAU, the patient is detoxified using a titration and tapering method [17].

**Table 1:** GHB withdrawal symptoms.

| **Mild – Moderate** | **Severe** |
| --- | --- |
| Fatigue/sleepiness | Hypertension |
| Insomnia | Tachycardia |
| Gloomy feeling | Hallucinations |
| Slow, sluggish feeling | Severe agitation |
| Tremor | Psychosis |
| Sweating | Delirium |
| Sudden cold/warm feeling | Epileptic seizures |
| Restlessness |  |
| Tensed, stressed feeling |  |
| Lively/unpleasant dreams |  |
| Fear |  |
| Yawning |  |
| Hungry |  |
| Muscle aches/twitches |  |
| Abdominal cramps |  |

##### Titration phase

Each patient will be administered 15mg baclofen every 4-6 hours, which is equivalent to 75mg per 24 hours in total. In addition to the scheduled baclofen dose 0,9-3ml pharmaceutical GHB (500mg/ml) will be added per gift (every 3 hours), depending on the severity of the withdrawal symptoms. In case of sedation, the baclofen dose is lowered with 2,5-5mg per gift. The titration phase continues until the patient is stable and experiences neither withdrawal nor sedation.

In case of lack of response to the baclofen and pharmaceutical GHB therapy after 5,5 hours, intolerable side effects, or any signs of severe withdrawal such as psychosis or delirium, the study will be discontinued, and a switch will be made to TAU.

##### Tapering phase

On the first day of the tapering phase, which starts when the patient has been stable on the baclofen and pharmaceutical GHB for one day, the pharmaceutical GHB (500mg/ml) will be tapered off with 0,6ml/gift/day (TAU). Once, the patient has completely tapered off the pharmaceutical GHB the baclofen dose will be reduced with 5-10mg per day, depending on the degree of withdrawal.

When the patient is stable for two days without pharmaceutical GHB and baclofen, the tapering phase and detoxification is completed. For an overview of the titration and tapering phase in the proof-of-concept study, see Figure 2.

**Figure 2.**
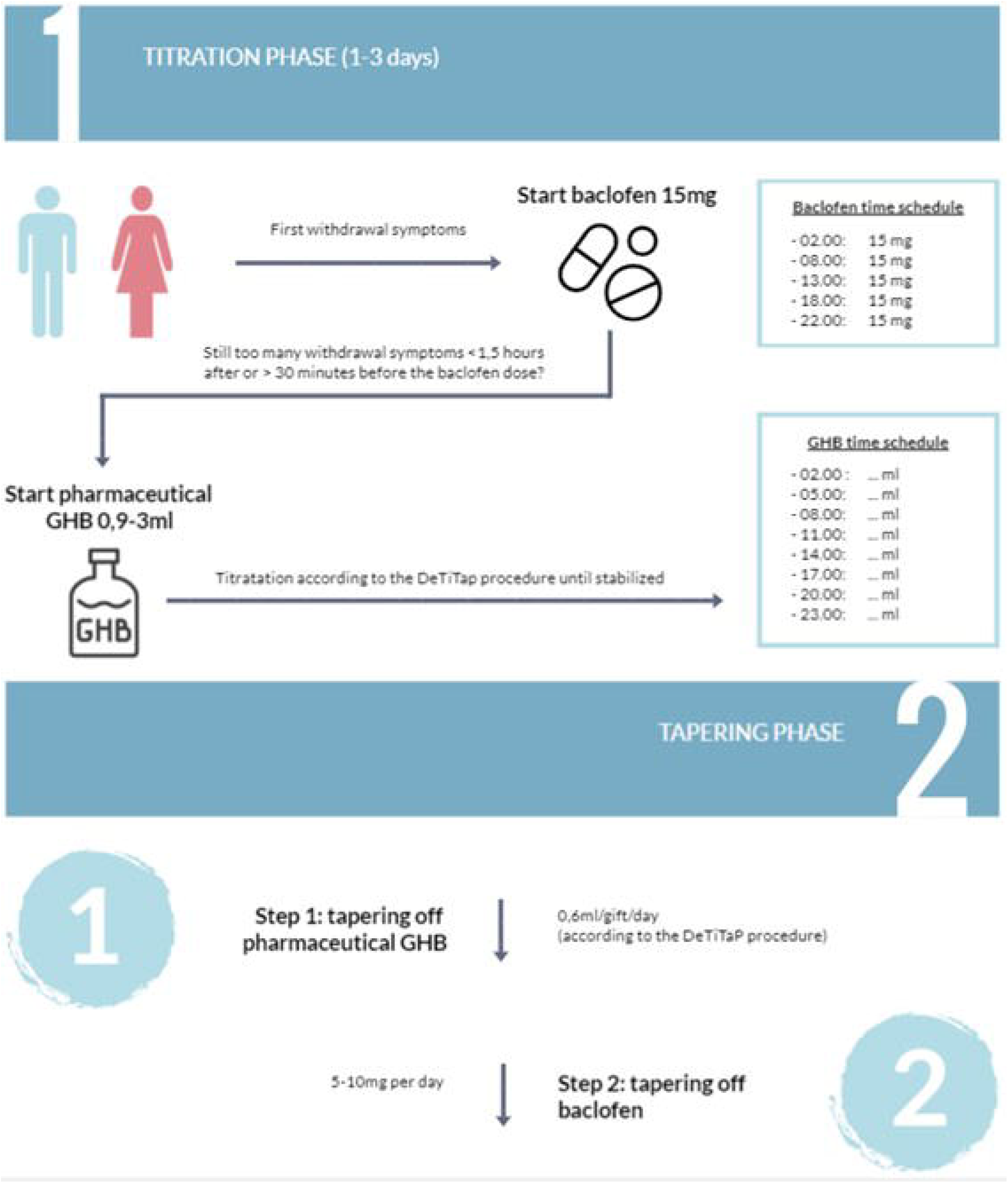
overview of titration and tapering phase of the proof-of-concept study (n= 10)

#### Baclofen dose-finding study

The protocol for the dose-finding study is identical to the proof-of-concept study and therefore also consists of a titration and tapering phase using the same method. To determine the adequate baclofen dosage during the titration phase, the baclofen dose will be increased in three steps. The first six patients will receive 17,5mg per 4-6 hours (87,5mg per day), the next six patients will be given 20mg per 4-6 hours (100mg per day), and the following six patients will be administered 22,5mg per 4-6 hours (112,5mg per day). For an overview, see figure 3.

**Figure 3:**
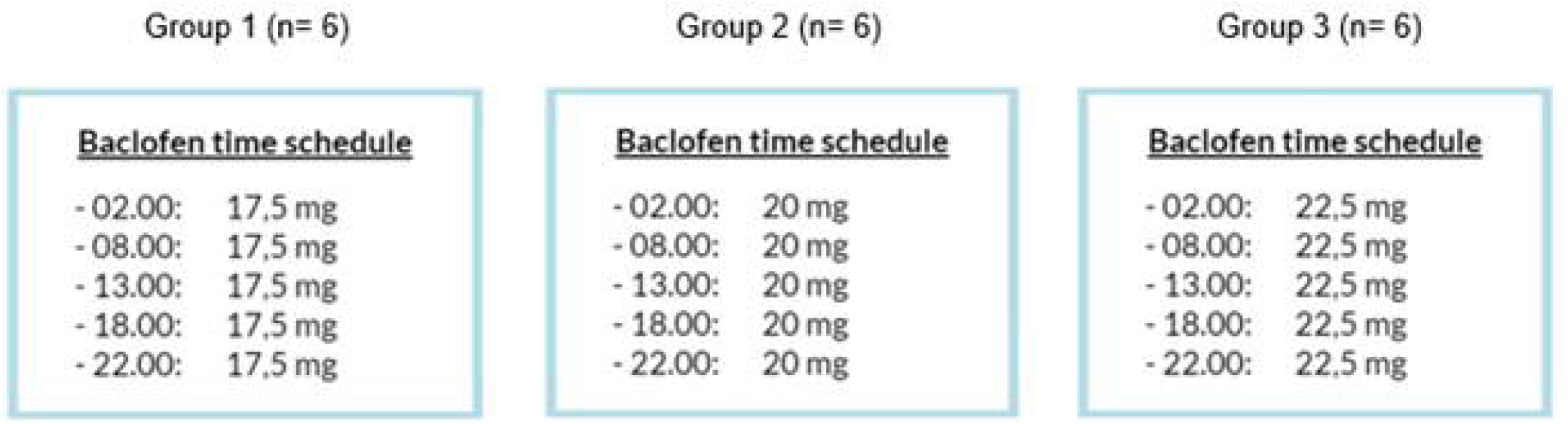
dose escalation steps during the proof-of-concept study (n= 18)

### Study parameters

The main study parameter is the dose of pharmaceutical GHB used by the participants, as recorded in the medical files. The secondary study parameters are subjective and objective withdrawal symptoms, craving and baclofen side effects.

Subjective and objective withdrawal severity will be assessed through the Subjective Withdrawal Scale (SWS) and Objective Withdrawal Scale (OWS). These scales are commonly used in GHB research [12,13,33,34] and are part of the regular detoxification procedure as described in the Dutch guideline for detoxification. The SWS consists of 33 items measuring withdrawal symptoms on a 5-point Likert scale ranging from 0 (not at all) till 4 (extremely). The OWS consists of 34 items and is filled out by trained health professionals, classifying withdrawal symptoms as ‘present’ or ‘absent’ based on their clinical observations.

Craving will be assessed by the Visual Analogue Scale (VAS). This is a commonly used single item 0-10 scale questionnaire in studies on SUD [13], which can easily be administered and is suitable for frequent and repeated measurements to detect rapid changes in a psychological state being assessed, such as the degree of craving [35].

Safety of baclofen will be monitored using a baclofen side effects questionnaire and by measuring vitals (e.g. blood pressure, heart rate and respiratory rate). The baclofen side effects questionnaire is based on the side effects of baclofen reported in the literature and has been used in earlier GHB studies from our group [23,36]. This questionnaire consists of 21 items with a five-point Likert scale ranging from 0 (never) till 4 (always), see Table 2 for the complete list.

**Table 2:**
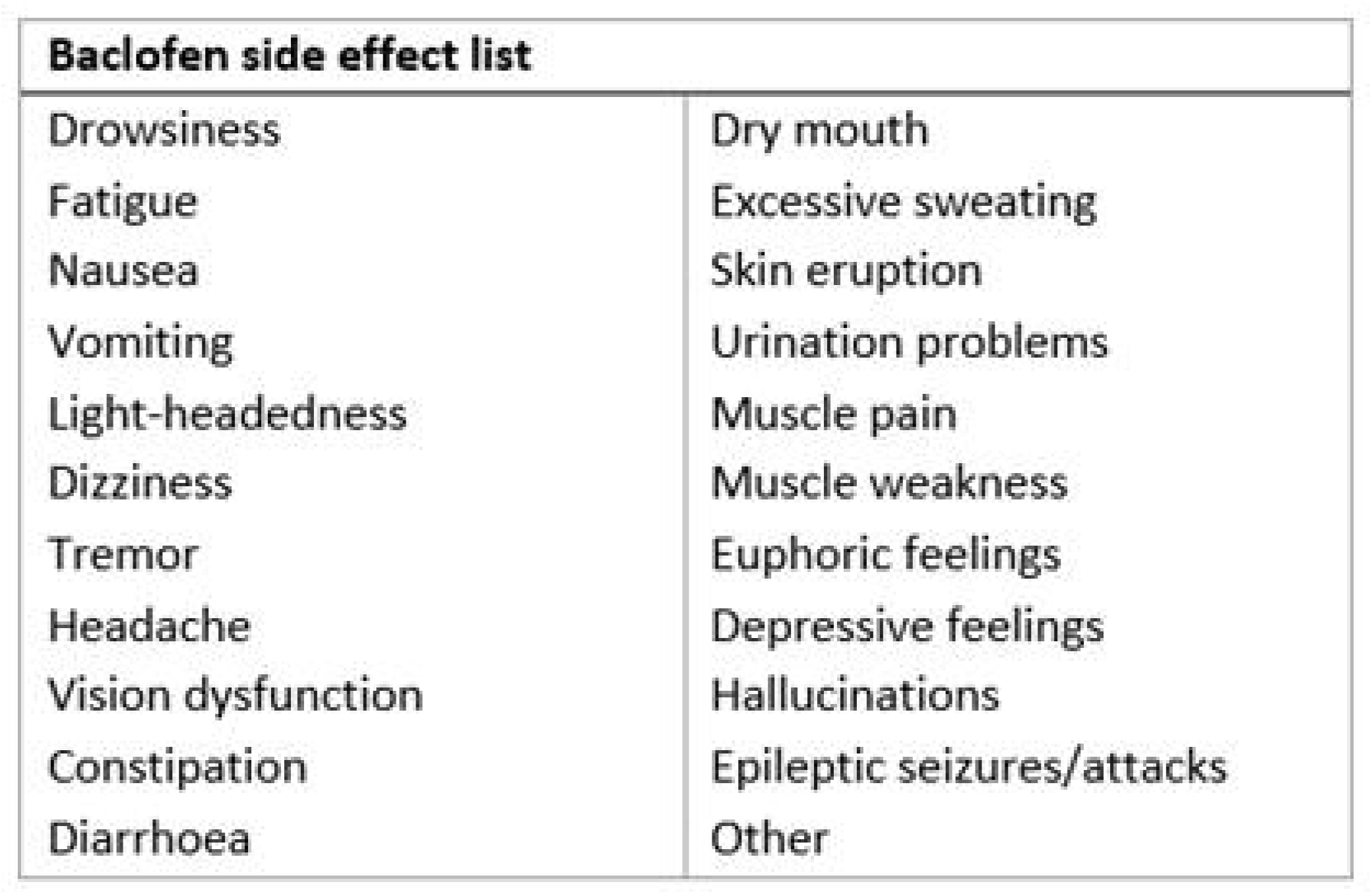
Baclofen side effects questionnaire list.

Other study parameters are general (socio)demographics and disease severity. General sociodemographic details and disease severity will be used to describe the included study population. The standard clinical assessment tool in Dutch addiction care treatment centres, which has proven to have good psychometric quality and good inter-rater reliability, is the Measurement of Addicts for Triage and Evaluation (MATE). The MATE measures the history, frequency and consequences of drug use, including medical, social and psychological problems [37]. For this study results from MATE, in which all substances/drugs used are described, will be included to assess current substance use (past 30 days), lifetime substance use, and the classification of substance dependence according to the DSM-IV [7].

Data about demographics (sex, date of birth, etc.) will be collected through self-report.

### Procedures

Patients will be recruited from the Radboud University Medical Centre and addiction treatment centre Novadic-Kentron, The Netherlands. Prior to admission to the clinic for inpatient detoxification, potential participants will be informed about the study and the off-label use of baclofen as detoxification method by the investigator. Subsequently, a medical doctor will screen potential participants for in- and exclusion criteria. Participants will provide written informed consent prior to inclusion in the study. Data on demographics and MATE are collected during the intake prior to admission to the clinic. Substance use in the past 30 days and medical and psychiatric history will be re-examined during the intake on the day of the clinical admission.

During the titration phase the SWS, OWS and VAS will be administered by trained nurses every 30 minutes before and after every GHB administration, to assess GHB withdrawal symptoms and craving during detoxification. Safety of baclofen will be monitored once a day using a baclofen side effects questionnaire and by measuring vitals (e.g. blood pressure, heart rate and respiratory rate).

During the tapering phase withdrawal symptoms and the degree of craving will be monitored three times a day, 30 minutes before a GHB administration, by a trained nurse using the SWS, OWS and VAS. Side effects of baclofen will be monitored once a week by a trained nurse using a baclofen side effects questionnaire.

### Data analysis

Data of all patients who completed the first day of the inpatient GHB detoxification will be included in the analysis. All of the following analysis will be done using IBM SPSS Statistics 29.

The pharmaceutical GHB and baclofen dosage (per gift during titration phase and average daily dosage during both titration- and tapering phase) and the duration of the titration and tapering phase will be presented quantitatively, as well as the subjective and objective withdrawal severity (SWS and OWS) and vitals.

In order to test whether addition of baclofen to TAU results in less pharmaceutical GHB during the titration phase (research objective 1.1), the pharmaceutical GHB dosage will be compared with the reference group using an independent samples t-test. The subjects in the reference group will be matched with the intervention group, based on the following factors (in order of priority): GHB dosage before admission to the clinic, comorbid drug use, psychiatric comorbidities, number of detoxifications in the past, gender and age.

To assess the safety of baclofen add-on therapy (research objective 1.2 and 2.3), the frequencies and descriptives of baclofen side effects will be presented. Occurrence of side effects and adverse events will be described descriptively.

Means, standard deviations and standard errors will be computed for withdrawal severity per day. A linear mixed model (LMM) analysis will be performed with withdrawal (SWS, OWS) and craving (VAS) scores as dependent variables and ‘days in admission’ (within-subjects variable) and ‘detoxification method’ (between-subjects variable) as fixed factors, to compare baclofen add-on therapy with TAU (research objective 2.1).

A LMM will be performed to assess the impact of different doses of baclofen on the need for pharmaceutical GHB, subjective withdrawal and craving during inpatient detoxification in patients with GUD (research objective 2.2). Pharmaceutical GHB dosage per gift during the titration phase and withdrawal (SWS, OWS) and craving (VAS) scores will be used as dependent variables and ‘baclofen dosage group’ (between-subjects variable) as fixed factors.

#### Sample size calculation

As no comparable studies have been conducted, an extensive sample size calculation could not be performed. For the proof-of-concept study ten patients will be included. For the dose-finding study another eighteen patients will be included. This is a reasonable sample size with which we expect to be able to answer our research questions. Furthermore, these findings can be used for future trials to adequately compute required sample sizes.

## Discussion

With current trial, we aim to investigate whether inpatient detoxification in patients with GUD can be improved with baclofen add-on treatment. The goals are to assess whether baclofen reduces the need for pharmaceutical GHB, to explore the optimal baclofen dosage and to assess the safety of baclofen as add-on therapy when used as add-on during detoxification with pharmaceutical GHB in patients with GUD.

One of the major gaps in literature, mainly consisting of case reports on baclofen addition, is that there is no consensus on the adequate baclofen dosage to manage GHB withdrawal symptoms. When used as add-on, most case reports report a daily dosage of 30-75mg baclofen divided into three doses in addition to the benzodiazepine regimen [21,28,29]. When used as stand-alone therapy to treat GHB withdrawal it is likely that higher baclofen doses are required [30]. Pre-clinical data combined with a number of case reports suggest that the use of baclofen is safe in supervised clinical settings up to 120mg per day [30,38]. In a review on the toxicity spectrum of baclofen in overdose [39], no severe clinical effects have been reported with baclofen doses up to 200mg. In this study, a maximum dose of 112,5mg is used to remain well within the margin of safety. However, the concurrent use of the central depressant GHB during detoxification warrants close monitoring of central sedative effects. In case of sedation, pharmaceutical GHB will be tapered off first. Given its short half-life, this is expected to swiftly result in a reduction of sedation. Since any increase in withdrawal symptoms will be counteracted by titrating pharmaceutical GHB as usual, the risk of under dosage is considered negligible.

In the current protocol, we titrate the baclofen dose faster than for registered indications, such as spasticity [38]. According to the pharmacotherapeutic compass, you normally start with 15mg/day and increase this to 75mg/day in twelve days [38]. However, due to the long-term excessive GHB use in our participants, it is likely that our study population has developed GABA tolerance allowing a faster titration schedule. This is in line with the reported high doses of benzodiazepines (GABA-A agonists) required to manage the GHB withdrawal symptoms [13,40] and further supported by the titration speed of baclofen in existing literature on GHB [22,25,27–29]. For example, in the case report of Habibian et al. (2019), 130mg baclofen was prescribed each day during the first two days, with a starting dose of 30mg and a dosage interval of 4 hours. Based on literature, expert opinion and clinical expertise, we believe that the rate of titration and tapering of baclofen in this study is suitable and safe for our study population.

Co-use of benzodiazepines is common in patients with GUD, as nearly 22-40% of the GUD population uses any kind of benzodiazepine [29,34]. Since benzodiazepines also act at the GABA-receptors, this might lead to undesirable interactions with both pharmaceutical GHB and baclofen. However, excluding all subjects with co-occurring benzodiazepine use would make our sample less representative for the GUD population. Therefore, only the use of benzodiazepines in doses above 30mg diazepam equivalent per day is excluded.

Based on current literature, we expect addition of 75mg baclofen per day will reduce the need for pharmaceutical GHB during GHB detoxification [21,25,27–29]. We hypothesize that higher doses up to 112,5mg will result in a greater reduction of pharmaceutical GHB. This will likely make GHB detoxification less burdensome for patients, and improve the overall feasibility of treatment. In addition, due to the long half-life of baclofen compared to pharmaceutical GHB, we expect less fluctuation in withdrawal symptoms during the day.

If the results of the current phase II trail are positive, a next step could be a replication phase II trial with a randomized blinded design or a phase III trial (therapeutic confirmative) to confirm the safety, feasibility and efficacy of baclofen add-on therapy in a larger study population. In addition, further dosage optimization and long-term safety of baclofen add-on therapy for inpatient GHB detoxification are important points of attention for subsequent studies.

## List of abbreviations

BZD: benzodiazepine
GABA: gamma-aminobutyric acid
GHB: gamma-hydroxybutyric acid
GUD: GHB use disorder
MATE: measurement of addicts for triage and evaluation
OWS: objective withdrawal scale
SUD: substance use disorder
SWS: subjective withdrawal scale
TAU: treatment as usual
VAS: Visual Analogue Scale

## Declarations

### Ethics approval and consent to participate

This study protocol is approved by the Medical Ethical Research Committee Oost-Nederland and Central Committee on Research Involving Human Subjects. The study is registered in the Clinical Trial Information System under EU CT-number: 2023-506167-34-02. All research is conducted in accordance with the Declaration of Helsinki, data protection laws and good clinical practice (GCP) as well as other relevant guidelines and regulations.

All patients will be asked to provide verbal and written informed consent for participation in this study. Minors or persons unable to provide informed consent will not be included.

### Consent for publication

Not applicable.

## Data availability statements

The datasets generated, used and/or analyzed during current study are available from the corresponding author upon reasonable request.

## Author Contributions

AW, HB, BD and AS designed the study. AW wrote the first draft of the manuscript. HB, BD and AS were involved in the supervision of the project. All authors critically revised the manuscript and gave final approval of the version to be submitted.

## Acknowledgements

The authors would like to thank prof. dr. Kees Kramers for his advisory role in pharmacology.

## References

1. Beurmanjer H, Asperslag EM, Oliemeulen L, Goudriaan AE, Jong CAJ De, Schellekens ASA, et al. A Qualitative Approach in Understanding Illness Perception and Treatment Needs in Patients with Gamma Hydroxybutyrate Use Disorder. Eur Addict Res. 2019;25(5).

2. Miotto K, Darakjian J, Basch J, Murray S, Zogg J, Rawson R. Gamma-hydroxybutyric acid: Patterns of use, effects and withdrawal. American Journal on Addictions. 2001;10(3):232–41.

3. Stein LAR, Lebeau R, Clair M, Martin R, Bryant M, Storti S, et al. A web-based study of gamma hydroxybutyrate (GHB): Patterns, experiences, and functions of use. American Journal on Addictions. 2011 Sep;20(1):30–9.

4. Sumnall HR, Woolfall K, Edwards S, Cole JC, Beynon CM. Use, function, and subjective experiences of gamma-hydroxybutyrate (GHB). Drug Alcohol Depend. 2008;92(1–3).

5. Kamal RM, Noorden MS Van, Franzek E, Dijkstra BAG, Loonen AJM, Jong CAJ De. The neurobiological mechanisms of gamma-hydroxybutyrate dependence and withdrawal and their clinical relevance: A review. Vol. 73, Neuropsychobiology. 2016.

6. Snead OC, Gibson KM. g-Hydroxybutyric Acid [Internet]. Vol. 352, n engl j med. 2005. Available from: https://www.nejm.org

7. American Psychiatric Association. American Psychiatric Association: Diagnostic and Statistical Manual of Mental Disorders Fifth Edition. Arlington. 2013.

8. Beurmanjer H, Kamal RM, de Jong CAJ, Dijkstra BAG, Schellekens AFA. Baclofen to Prevent Relapse in Gamma-Hydroxybutyrate (GHB)-Dependent Patients: A Multicentre, Open-Label, Non-Randomized, Controlled Trial. CNS Drugs. 2018 Sep;32(5):437–42.

9. Tay E, Lo WKW, Murnion B. Current Insights on the Impact of Gamma-Hydroxybutyrate (GHB) Abuse. Subst Abuse Rehabil. 2022 Sep;Volume 13:13–23.

10. van Noorden MS, Mol T, Wisselink J, Kuijpers W, Dijkstra BAG. Treatment consumption and treatment re-enrollment in GHB-dependent patients in The Netherlands. Drug Alcohol Depend. 2017;176.

11. McDonough M, Kennedy N, Glasper A, Bearn J. Clinical features and management of gamma-hydroxybutyrate (GHB) withdrawal: A review. Vol. 75, Drug and Alcohol Dependence. Elsevier Ireland Ltd; 2004. p. 3–9.

12. Wolf CJH, Beurmanjer H, Dijkstra BAG, Geerlings AC, Spoelder M, Homberg JR, et al. Characterization of the ghb withdrawal syndrome. J Clin Med. 2021 Sep;10(11).

13. Beurmanjer H, Luykx JJ, Wilde B De, van Rompaey K, Buwalda VJA, Jong CAJ De, et al. Tapering with Pharmaceutical GHB or Benzodiazepines for Detoxification in GHB-Dependent Patients: A Matched-Subject Observational Study of Treatment-as-Usual in Belgium and The Netherlands. CNS Drugs. 2020;34(6).

14. Hu R, Banerjee P, Snead III O. Regulation of γ-aminobutyric acid (GABA) release in cerebral cortex in the γ-hydroxybutyric acid (GHB) model of absence seizures in rat [Internet]. Vol. 39, Neuropharmacology. 2000. Available from: https://www.elsevier.com/locate/neuropharm

15. Carter LP, Koek W, France CP. Behavioral analyses of GHB: Receptor mechanisms. Vol. 121, Pharmacology and Therapeutics. 2009. p. 100–14.

16. Liechti ME, Quednow BB, Liakoni E, Dornbierer D, Von Rotz R, Gachet MS, et al. Pharmacokinetics and pharmacodynamics of γ-hydroxybutyrate in healthy subjects. Br J Clin Pharmacol. 2016 May 1;81(5):980–8.

17. Dijkstra BAG, Oort MMHJ, van Schellekens AFA, Haan HA, de Jong CAJde. Richtlijn detoxificatie van psychoactieve middelen: Verantwoord ambulant of intramuraal detoxificeren [Internet]. 2017 [cited 2023 Jan 10]. Available from: https://hdl.handle.net/2066/174019

18. Kamal RM, van Noorden MS, Wannet W, Beurmanjer H, Dijkstra BAG, Schellekens A. Pharmacological Treatment in γ-Hydroxybutyrate (GHB) and γ-Butyrolactone (GBL) Dependence: Detoxification and Relapse Prevention. Vol. 31, CNS Drugs. 2017.

19. van Noorden MS, van Dongen LCAM, Zitman FG, Vergouwen T (A)C M. Gamma-hydroxybutyrate withdrawal syndrome: dangerous but not well-known. Gen Hosp Psychiatry. 2009 Jul;31(4):394–6.

20. Lingford-Hughes A, Patel Y, Bowden-Jones O, Crawford MJ, Dargan PI, Gordon F, et al. Improving GHB withdrawal with baclofen: Study protocol for a feasibility study for a randomised controlled trial. Trials. 2016;17(1).

21. Agabio R, Preti A, Gessa GL. Efficacy and tolerability of baclofen in substance use disorders: A systematic review. Vol. 19, European Addiction Research. 2013. p. 325–45.

22. Floyd CN, Wood DM, Dargan PI. Baclofen in gamma-hydroxybutyrate withdrawal: patterns of use and online availability. Eur J Clin Pharmacol. 2018;74(3).

23. Kamal RM, Schellekens A, Jong CAJ De, Dijkstra BAG. Baclofen as relapse prevention in the treatment of Gamma-Hydroxybutyrate (GHB) dependence: An open label study. BMC Psychiatry. 2015;15(1).

24. Crunelli V, Emri Z, Leresche N. Unravelling the brain targets of γ-hydroxybutyric acid. 2006.

25. LeTourneau JL, Hagg DS, Smith SM. Baclofen and gamma-hydroxybutyrate withdrawal. Neurocrit Care. 2008 Sep;8(3):430–3.

26. Fattore L, Cossu G, Martellotta MC, Deiana S, Fratta W. Baclofen antagonises intravenous self-administration of γ-hydroxybutyric acid in mice. Neuroreport. 2001;12(10).

27. Bell J, Collins R. Gamma-butyrolactone (GBL) dependence and withdrawal. Addiction. 2011 Feb;106(2):442–7.

28. Lai W, Raposa JT, Parlapalli R. Treatment of Poorly Responsive Gamma-Hydroxybutyrate Withdrawal With Baclofen: A Case Report. Cureus. 2022 Sep;

29. Siefried KJ, Freeman G, Roberts DM, Lindsey R, Rodgers C, Ezard N, et al. Inpatient GHB withdrawal management in an inner-city hospital in Sydney, Australia: a retrospective medical record review. Psychopharmacology (Berl). 2022 Jan 1;

30. Habibian S, Ahamad K, McLean M, Socias ME. Successful Management of Gamma-hydroxybutyrate (GHB)Withdrawal Using Baclofen as a Standalone Therapy: A Case Report. J Addict Med. 2019;13(5).

31. Müller CA, Geisel O, Pelz P, Higl V, Krüger J, Stickel A, et al. High-dose baclofen for the treatment of alcohol dependence (BACLAD study): A randomized, placebo-controlled trial. European Neuropsychopharmacology. 2015 Aug 1;25(8):1167–77.

32. Reynaud M, Aubin HJ, Trinquet F, Zakine B, Dano C, Dematteis M, et al. A randomized, placebo-controlled study of high-dose baclofen in alcohol-dependent patients-The ALPADIR study. Alcohol and Alcoholism. 2017 Jul 1;52(4):439–46.

33. Beurmanjer H. When the party is over. Adressing clinical challenges in patients with GHB use disorders. 2021;

34. Dijkstra BAG, Kamal R, van Noorden MS, de Haan H, Loonen AJM, Jong CAJ De. Detoxification with titration and tapering in gamma-hydroxybutyrate (GHB) dependent patients: The Dutch GHB monitor project. Drug Alcohol Depend. 2017;170.

35. Rosenberg H. Clinical and laboratory assessment of the subjective experience of drug craving. Vol. 29, Clinical Psychology Review. 2009.

36. Kamal RM, Loonen AJM, Dijkstra BAG, De Jong CAJ. Baclofen as relapse prevention in the treatment of gamma-hydroxybutyrate dependence. J Clin Psychopharmacol. 2015 Jun 13;35(3):313–8.

37. Schippers GM, Broekman TG, Buchholz A, Koeter MWJ, Brink W Van Den. Measurements in the Addictions for Triage and Evaluation (MATE): An instrument based on the World Health Organization family of international classifications. Addiction. 2010;105(5).

38. Zorginstituut Nederland. Farmacotherapeutisch Kompas. Retrieved from https://farmacotherapeutischkompas.nl. Farmacotherapeutisch Kompas..

39. Leung NY, Whyte IM, Isbister GK. Baclofen overdose: Defining the spectrum of toxicity. EMA -Emergency Medicine Australasia. 2006 Feb;18(1):77–82.

40. Karila L, Angerville B, Benyamina A, Billieux J. Pharmacological Treatment of GHB Withdrawal Syndrome. Vol. 11, Current Addiction Reports. Springer Science and Business Media Deutschland GmbH; 2024. p. 163–71.

